# Evaluation of four interventions using behavioural economics insights to increase demand for voluntary medical male circumcision in South Africa through the MoyaApp: A quasi-experimental study

**DOI:** 10.1101/2024.01.18.24301032

**Authors:** Preethi Mistri, Silviu Tomescu, Simamkele Bokolo, Alexandra De Nooy, Pedro T. Pisa, Skye Grove, Laura Schmucker, Candice Chetty-Makkan, Lawrence Long, Alison Buttenheim, Brendan Maughan-Brown

## Abstract

**Background:** While voluntary medical male circumcision (VMMC) reduces the risk of HIV transmission by 60%, circumcision coverage falls short of the UNAIDS 90% target. We investigated whether behaviourally informed message framing increased demand for VMMC.

**Setting:** Adult users of the MoyaApp, a data free application in South Africa, who viewed a form designed to generate interest in VMMC during August-November 2022.

**Methods:** A quasi-experimental study was conducted to evaluate four MoyaApp VMMC intervention forms against the Standard of Care (SOC) form. All forms enabled users to provide contact details for follow-up engagement by a call centre. The primary outcome was the proportion of forms submitted. Secondary outcomes included successful contact with the user, VMMC bookings/referrals and confirmed circumcision. Multivariable ordinary least-squares regression was used for the analysis.

**Results:** MoyaApp VMMC form viewers totalled 118,337 of which 6% submitted a form. Foot-in-the-Door form viewers were more likely (+1.3 percentage points, p<0.01) to submit a form compared to the SOC group (6.3%). Active Choice (-1.1 percentage points, p<0.01) and Reserved for You (-0.05 percentage points, p<0.05) form viewers were less likely to submit a form compared to SOC. Users submitting on Foot-in-the-Door were less likely to be booked/referred compared to SOC (-5 percentage points, p<0.05). There were no differences between the intervention and SOC forms for successful contact and circumcisions.

**Conclusions:** Message framing using behavioural insights was able to nudge men to engage with VMMC services. However, more work is needed to understand how to convert initial interest into bookings and circumcisions.

**Trial registration:**

- South African Clinical Trials Registry DOH-27-062022-7811
- Pan-African Clinical Trials Registry PACTR202112699416418

## Introduction

Voluntary medical male circumcision (VMMC) is effective in reducing the risk of HIV transmission (female-to-male by approximately 60%^1^ and male-to-male by approximately 23%^2^) and offers lifelong risk reduction for HIV transmission^3^. The WHO and UNAIDS recommend VMMC as a core HIV-prevention strategy, setting a target of 90% coverage for VMMC in high-priority countries, including South Africa^4^. Between 2008 and 2019, 26.8 million men and boys were circumcised in sub-Saharan Africa and an estimated 340,000 new HIV infections were averted by 2019^5^. While more than 4.4 million men in South Africa were circumcised during this time, the country still falls short of the 90% target with only 62.5% of men between 15-49 years old reported to be circumcised in 2022^6^.

Multiple barriers to VMMC include fear of the pain of the procedure, concerns around the abstinence period post-medical circumcision, stigma around HIV testing (which is required pre- circumcision), perceived low HIV risk, loss of income as a result of missed work and the belief that VMMC is not appropriate for older males^7,8^. Additionally, men are less likely to engage in HIV health services than women, which may be exacerbated by concerns regarding confidentiality, perceptions of compromised masculinity, and stigma^1,9^. This adds to the challenge of motivating VMMC uptake.

Extensive demand creation strategies have been central to the scale-up of VMMC in priority countries^8,10,11^. These included mass media and community mobilisation strategies often using behaviour change communication materials to improve knowledge and awareness of medical circumcision through radio broadcasting, social media, vehicle advertising, billboards, distribution of informational leaflets, community events, engagement of community leaders, traditional healers, male-friendly clinics, and school education systems^8,10–12^. Frameworks built on data from VMMC programme implementation and market research indicate that demand-generation activities should include innovative approaches to motivate men to accept services and address gaps in VMMC engagement^8,13^. These include tailoring messages to men’s stage of behaviour change (i.e., behaviour change can be a multi-stage process with varying levels of awareness, readiness, intentions, etc., at each stage), including benefits other than just HIV prevention in messaging, promoting positive social norms and addressing individual and environmental barriers^8,13^.

Decisions may be influenced based on how they are framed or presented^14^ and also by psychological, behavioural, emotional, social and contextual factors^15^. Messages can be constructed to address specific behavioural barriers to overcome a potential intention-action gap or increase intention^13,16^. We used message framing leveraging behavioural economics principles of social norms, the endowment effect, exclusivity, loss aversion, the foot-in-the-door technique and active choice to increase demand for VMMC in South Africa.

## Methods

### Study Design and Population

This was a five-arm quasi-experimental, non-randomised group study conducted 8 August to 21 November 2022 across nine provinces in South Africa. The target sample included adults (≥18 years old) registered on the MoyaApp who accessed a VMMC form during the study period.

The MoyaApp is a data free app which is available on the four main mobile network providers in South Africa with approximately 4 million daily users and 6.5 million monthly users accessing the platform^17^. A VMMC icon in the MoyaApp health section directs individuals to a VMMC form where they can submit contact details as an expression of interest for a call back by programme counsellors. The VMMC form on the MoyaApp and related VMMC demand generation activities are implemented by Right to Care (RTC), a healthcare organisation supporting HIV prevention and treatment programmes in South Africa.

The VMMC form is created in Microsoft Forms and manually uploaded to the MoyaApp platform, which can only host one form at any given time. The study evaluated four new intervention forms against the Standard of Care (SOC) form (five forms overall). The forms were allocated to different blocks of time in the study period. The 15-week study period was divided into 45 blocks of time, with forms changed three times a week, at approximately 8am (SAST) every Monday, Wednesday and Friday. It was operationally feasible to make three form rotations per week within the VMMC programme. Forms were allocated to the 45 study blocks in a predetermined sequence, with all five forms being used consecutively and the same sequence being repeated (Supplementary Table S1).

The initial sequence of the five forms was randomised. Each form was used nine times (i.e. 45 study blocks divided by five forms) on the MoyaApp at roughly equal timepoints across the study period, which aimed to account for potential temporal influences on app usage and circumcision demand.

Submission of a form generated an automated email to the call centre, triggering counsellors to contact individuals telephonically. Counsellors at the call centre provided information on VMMC, answered questions, and then either scheduled an appointment for the procedure or referred interested clients to other service providers. Counsellors made multiple attempts to reach clients using both primary and secondary (if provided) contact numbers.

### Control group: Standard of Care form

The SOC form contained a banner identifying the medical circumcision program, an image and text with fewer than 60 words. The SOC provided general information and health benefits of VMMC. The form contained fields to capture name, surname, contact number, alternate number and age category (≥18 years old or under 18 years). See Box 1 for a detailed description of the SOC and intervention forms.

**Box 1.**
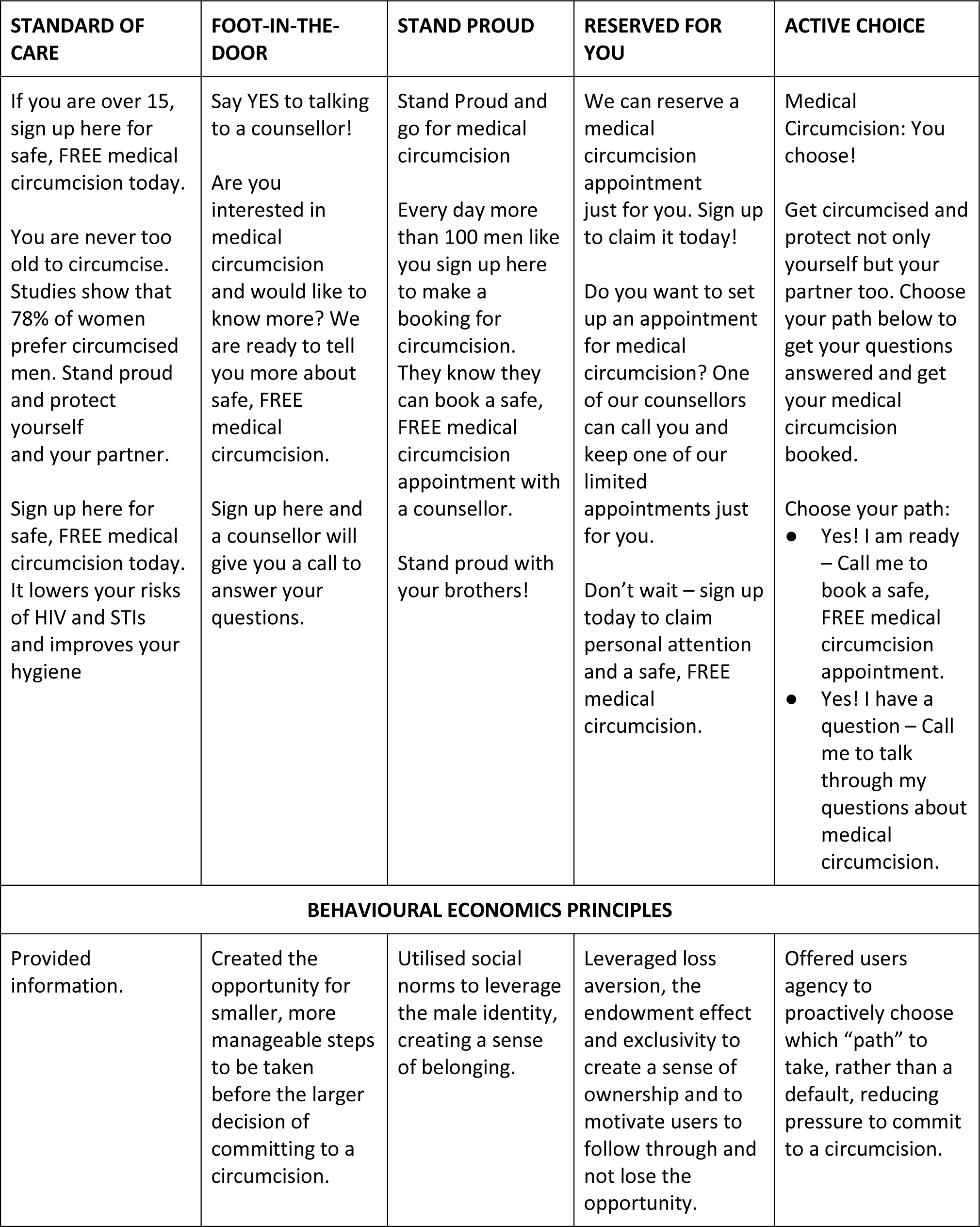
Form title and messaging.

Box 1 Form title and messaging
| STANDARD OF CARE | FOOT-IN-THE-DOOR | STAND PROUD | RESERVED FOR YOU | ACTIVE CHOICE |
| --- | --- | --- | --- | --- |
| If you are over 15, sign up here for safe, FREE medical circumcision today.<br>You are never too old to circumcise. Studies show that 78% of women prefer circumcised men. Stand proud and protect yourself and your partner.<br>Sign up here for safe, FREE medical circumcision today. It lowers your risks of HIV and STIs and improves your hygiene | Say YES to talking to a counsellor!<br>Are you interested in medical circumcision and would like to know more? We are ready to tell you more about safe, FREE medical circumcision.<br>Sign up here and a counsellor will give you a call to answer your questions. | Stand Proud and go for medical circumcision<br>Every day more than 100 men like you sign up here to make a booking for circumcision. They know they can book a safe, FREE medical circumcision appointment with a counsellor.<br>Stand proud with your brothers! | We can reserve a medical circumcision appointment just for you. Sign up to claim it today!<br>Do you want to set up an appointment for medical circumcision? One of our counsellors can call you and keep one of our limited appointments just for you.<br>Don't wait – sign up today to claim personal attention and a safe, FREE medical circumcision. | Medical Circumcision: You choose!<br>Get circumcised and protect not only yourself but your partner too. Choose your path below to get your questions answered and get your medical circumcision booked.<br>Choose your path:<br>● Yes! I am ready – Call me to book a safe, FREE medical circumcision appointment.<br>● Yes! I have a question – Call me to talk through my questions about medical circumcision. |
| BEHAVIOURAL ECONOMICS PRINCIPLES |  |  |  |  |
| Provided information. | Created the opportunity for smaller, more manageable steps to be taken before the larger decision of committing to a circumcision. | Utilised social norms to leverage the male identity, creating a sense of belonging. | Leveraged loss aversion, the endowment effect and exclusivity to create a sense of ownership and to motivate users to follow through and not lose the opportunity. | Offered users agency to proactively choose which “path” to take, rather than a default, reducing pressure to commit to a circumcision. |

### Interventions

The intervention forms were co-designed with the implementing partner including the VMMC programme strategic, data, technical and operational staff who provided insights on behavioural barriers to VMMC services. Behavioural economics principles leveraged included social norms, loss aversion, the endowment effect, exclusivity, the foot-in-the-door persuasion technique and active choice. Message framing made the steps towards VMMC feel easy and low-cost.

Six forms were initially designed, prototyped, and refined, before the four most promising forms were selected to be evaluated in the study. Intervention forms included the same fields for contact details as the SOC form. The call centre followed standard programmatic procedures across all forms following submissions and during contact and engagement with users on the VMMC programme.

#### Intervention Form 1: Foot-in- the-Door

The foot-in-the-door persuasion technique in which individuals are asked to take a small initial step towards a big decision^18–20^, was used to frame form submission and telephonic engagement with a counsellor as a small, manageable step before the larger decision to commit to VMMC.

#### Intervention Form 2: Stand Proud

The Stand Proud form was designed on the premise that individual behaviour is greatly influenced by what people see or hear of others doing. Social norms - the informal rules of beliefs, attitudes, and behaviours that are considered acceptable in a particular society or social group^21^ - have been leveraged to influence a range of behaviours^22,23^.

#### Intervention Form 3: Reserved for You

If people understand that something is reserved for them it creates a sense of exclusivity and access to a “coveted opportunity” which amplifies the perceived value of the benefit on offer^24^. Coupled with this implied selectivity, this form utilised loss aversion (an individual’s tendency to prefer avoiding losses to acquiring similar gains^25^) and the endowment effect, to personalise the appointment booking process and address barriers related to perceptions of healthcare-related inconveniences.

#### Intervention Form 4: Active Choice

This form provided users agency to choose a path, and to reduce pressure to commit to VMMC by requesting them to proactively make a choice rather than having a default choice already selected^26^. The decision architecture catered for people who may have already decided to go ahead with circumcision and were ready to make a booking; as well as those people who still had questions about VMMC and needed further engagement and information before making the decision.

### Data and outcomes

Routine administrative data were used for this study, with no primary data collection. The primary study outcome was the proportion of MoyaApp users viewing a VMMC submission form who then submitted the form. For the primary outcome, the service provider (MoyaApp) provided data on the number of MoyaApp users who viewed a VMMC form in each of the 45 study blocks (i.e., between the time a form was uploaded to when it was changed). Data was retrieved from the RTC VMMC programme for the number of form submissions, call centre contact attempts and outcomes, bookings or referrals and circumcisions. A unique study identification number (ID) was created for every individual who submitted a form, with repeat form submissions and call centre outcomes identified by cell phone number and assigned to the relevant ID. Data for attending a clinic visit for the VMMC procedure was retrieved from the RightMax Lynx electronic data system, which was linked to the VMMC programme data using national identity numbers.

Three binary variables were created as secondary outcomes that were based on the key steps that individuals took along the VMMC journey: 1) Contacted (=1) by the call centre captured whether the call centre was able to speak with someone telephonically using the contact details provided on the submitted VMMC form; 2) Booked/referred for a VMMC appointment (=1) identified individuals whom the call centre agent spoke with and who were either booked directly for a VMMC appointment or referred to another service provider (which is the protocol for clients residing outside of regions where RTC operates); and 3) Circumcised (=1) identified individuals who were recorded in the clinic system as having attended their VMMC appointment and been circumcised.

### Statistical analysis

For each outcome, Chi-squared tests were first used to assess the difference in proportions between the SOC form and each of the intervention forms. For ease of interpretation, linear probability models (ordinary least-squares regression) were used to estimate the effect of each intervention form relative to the SOC, with adjustment to account for the clustering of observations within each of the 45 study blocks, and controlling for potential study week and day of week (Monday, Wednesday or Friday) effects^27^. For robustness, we also estimated logistic regression models. Analyses were conducted in Stata V.17^28^.

#### Form submissions (primary outcome)

Data were excluded (n=26) when a form was submitted during the period in which the form was being changed from one study group to another, and it was not possible to know which form was viewed by the individual.

#### Call centre contact, VMMC booking and circumcision (secondary outcomes)

The impact of the intervention on the secondary outcomes was assessed among the sub-sample of individuals who both submitted a form and were included in the list of individuals for subsequent contact.

An administrative data system error resulted in a proportion of individuals who submitted a form (2498/6652) not being included in the list (i.e., no contact attempts were made based on these forms). The analysis of the intervention effects on the secondary outcomes was designed as an individual-level analysis. Individuals were assigned to a study group based on the first form they submitted, with the rationale that exposure to the first form could influence all subsequent decisions. Additional regression analysis was conducted for the secondary outcomes to assess for robustness to the inclusion of an additional binary control variable which captured whether an individual submitted multiple forms (=1) or not (=0). It is possible that the call centre could have made more attempts to contact these individuals if the details submitted were not recognised as duplicate.

### Ethical considerations

The study was approved by the Human Research Ethics Committee (Medical) of the University of the Witwatersrand, and the University of Pennsylvania, and Boston University institutional review boards. A waiver of informed consent was granted for the study which utilised anonymised administrative data.

## Results

Overall, there were 118,337 MoyaApp VMMC form viewers 18 years and older during the study period. The total number of form viewers included in the primary analysis was 24,459 for SOC, 22,351 for Foot-in-the-Door, 23,195 for Stand Proud, 24,178 for Reserved for You, and 24,154 for Active Choice (Figure 1).

**Figure 1.**
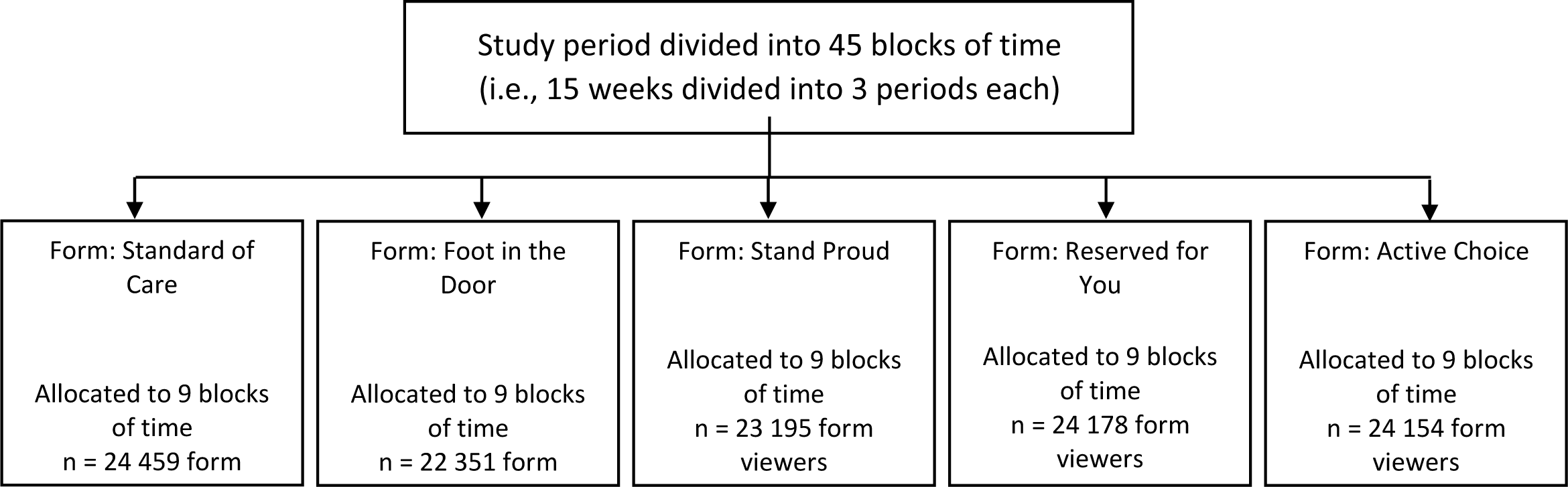
Flow of study participants for MoyaApp voluntary medical male circumcision form viewers 18 years and older

### Form submissions (primary analysis)

Out of 118,337 form viewers across all study blocks, 7089 (6%) submitted a VMMC form with 6.3% submitting on the SOC form. A higher proportion submitted the Foot-in-the-Door form (7.4%) than the SOC form (p<0.01). The proportion submitting in all other forms was lower than the SOC form: Stand Proud (5.8%, p<0.05), Reserved for You (5.6%, p<0.01) and Active Choice (5.0%, p<0.01) (Figure 2A).

**Figure 2.**
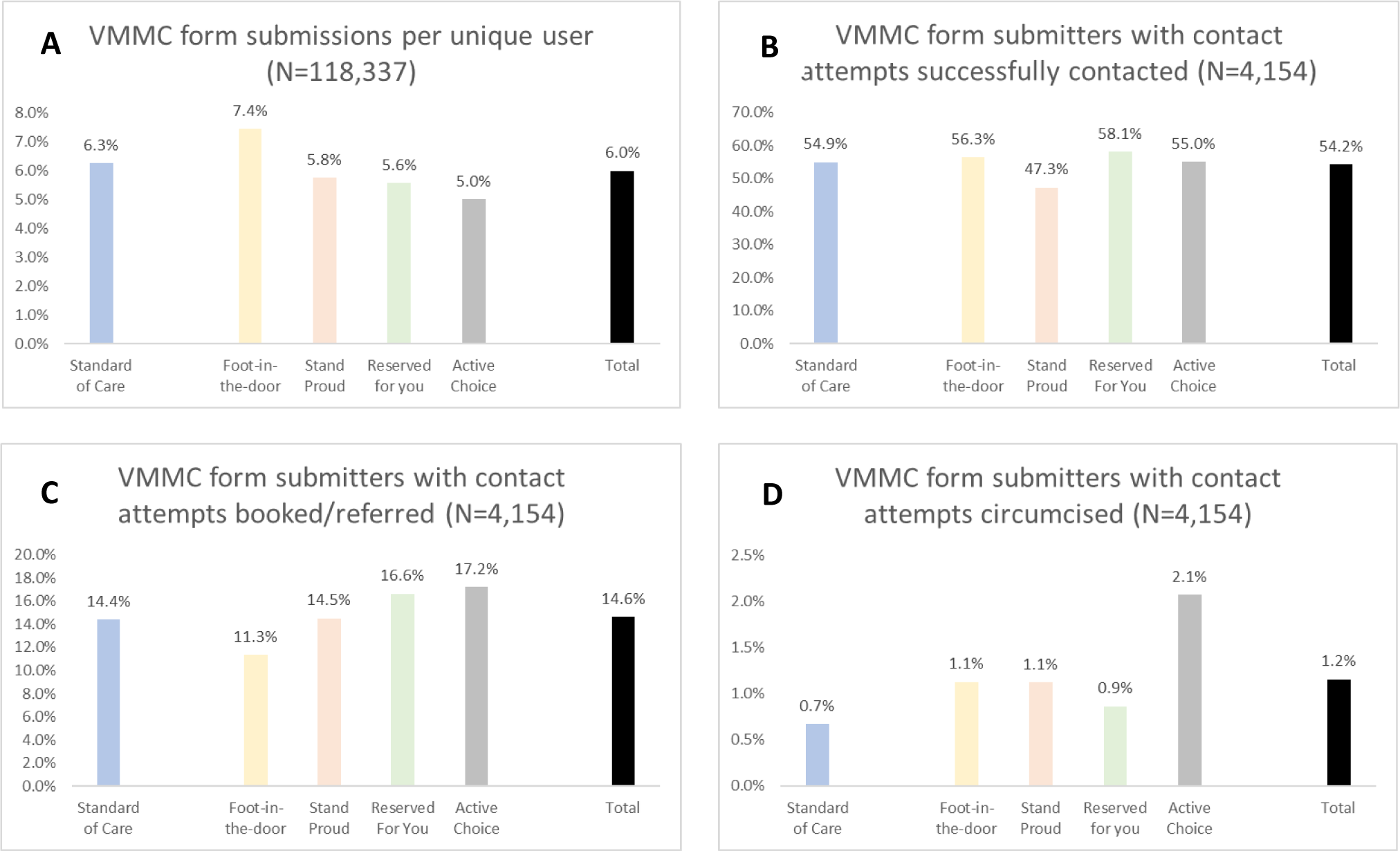
Proportion of MoyaApp users submitting forms and proportions of form submitters with contact attempts successfully contacts, booked/referred and circumcised

Ordinary least squares regression results, adjusted for potential confounders (Table 1, Model 2) showed that viewers of the Foot-in-the-Door form were more likely (+1.3 percentage points, p<0.01) to submit than those who viewed the SOC form. Contrastingly, compared to the SOC form, submissions were lower on the Active Choice (-1.1 percentage points, p<0.01) and Reserved for You forms (-0.05 percentage points, p<0.05). Associations with day of the week and study week number controls can be seen in Supplementary Table S2. Results were substantively similar in logistic regression models (Supplementary Table S3).

**Table 1.**
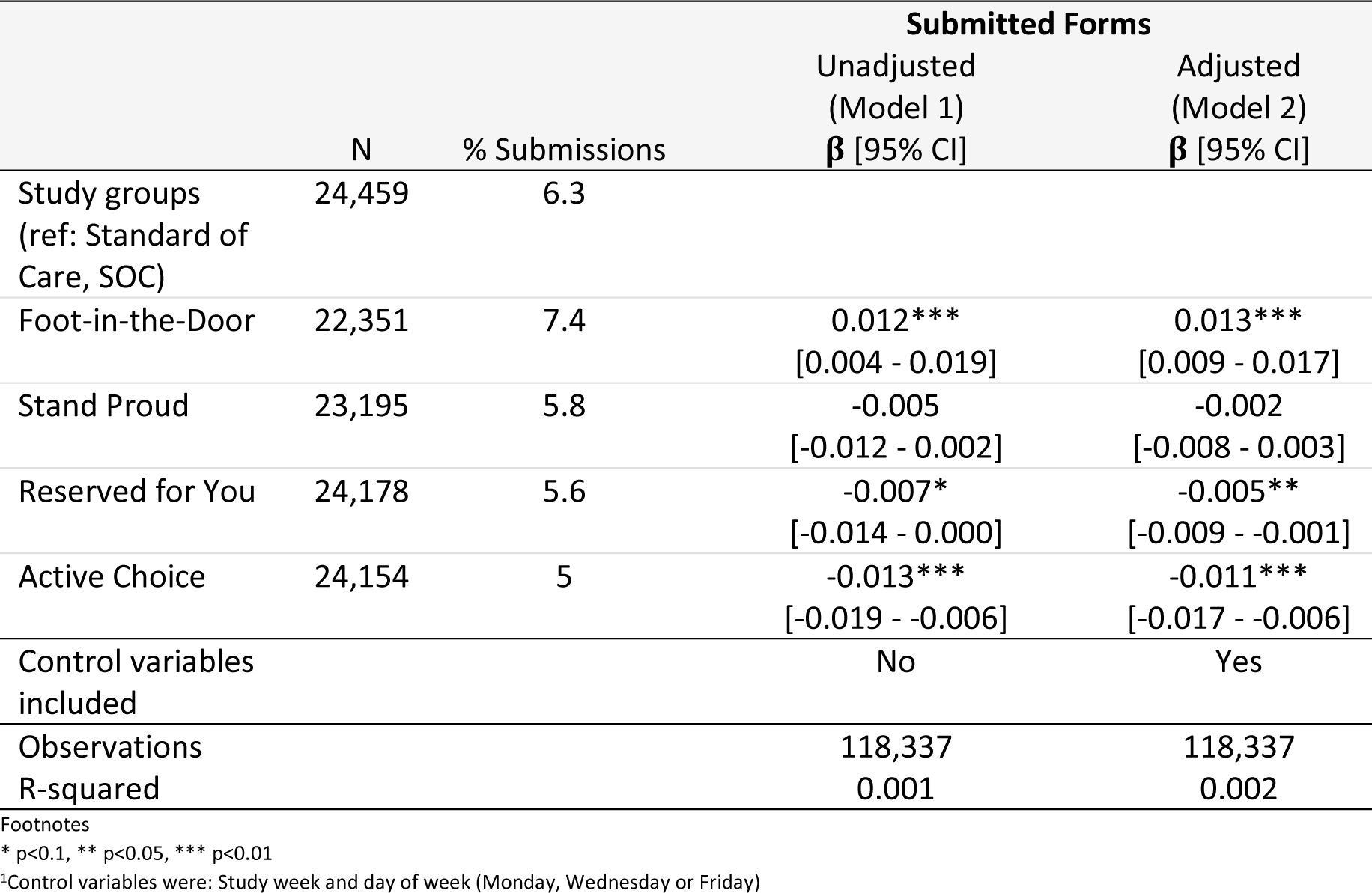
Ordinary least squares regression models assessing the association between VMMC forms submitted across five arms by adults who viewed the forms during August and November 2022 in South Africa (N=118,337)

### Call centre contact, VMMC booking and circumcision (secondary analysis)

The secondary analysis included the sub-sample of 4,154 individuals who submitted a form and their form was included in the list of individuals to be contacted by the call centre. Additional analyses (Supplementary Table S4) showed that the SOC form was excluded from the contact list more than the intervention forms.

### Contacted

Of the adult users submitting a form and who were on the contact list, 54.2% were successfully contacted. More users were contacted after submitting a Reserved for You (58.1%) and Foot-in-the-Door (56.3%) form compared to SOC (54.9%), Active Choice about the same (55.0%) and Stand Proud worse (47.3%) (Figure 2B).

Regression results (Table 2, Model 2) showed that the difference in successful contacts between the SOC and any of the intervention forms was not statistically different after accounting for clustering and adjusting for potential confounders. Full model results for all secondary analyses are provided in Supplementary Table S5.

**Table 2.**
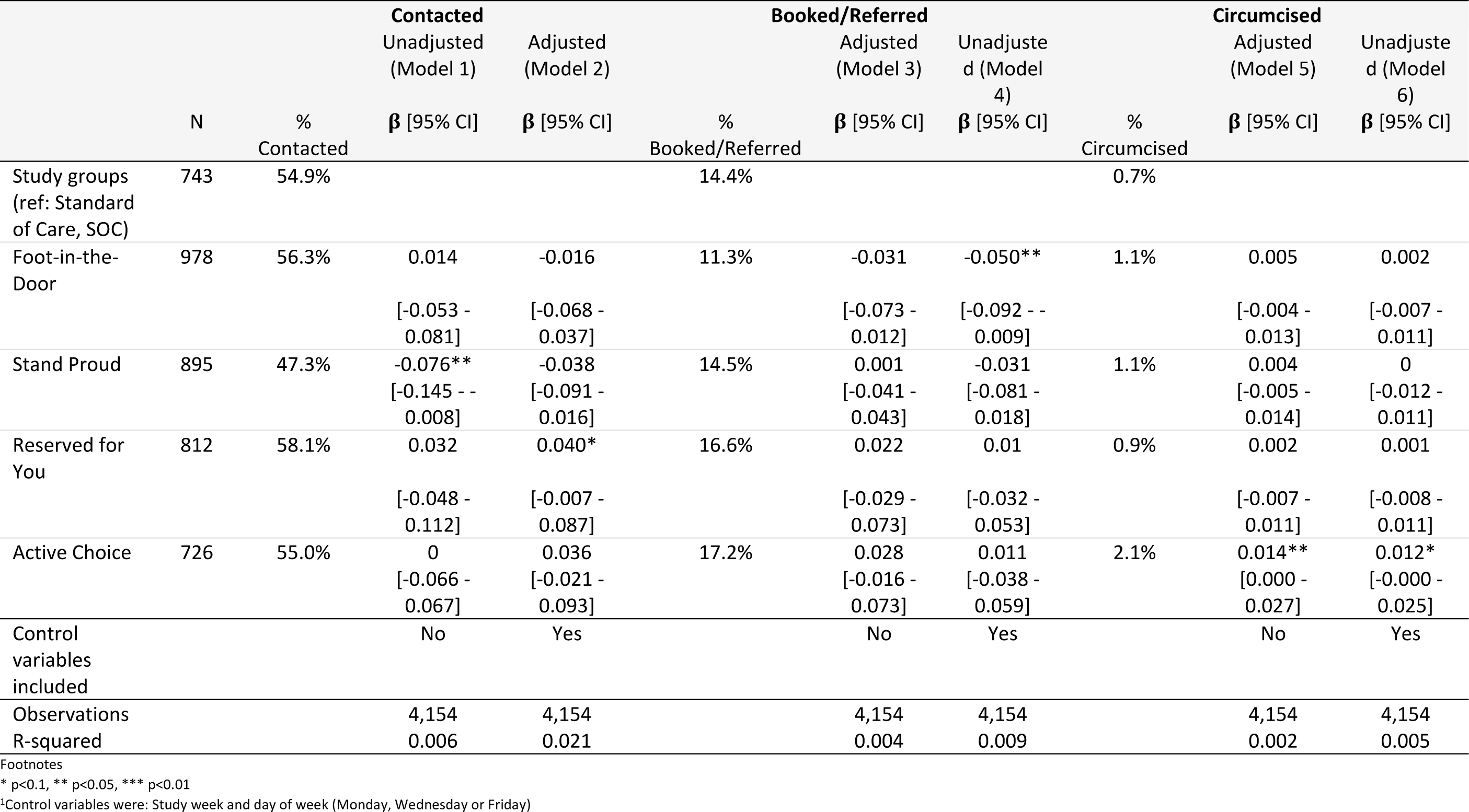
Ordinary least squares regression models of the impact of the study forms on secondary study outcomes: successful contact by the call centre, booking/referral complete, circumcision complete

### Booked/Referred

Among form submitters, 14.6% were booked or referred for VMMC. Compared to SOC (14.4%), more bookings were made among individuals who submitted on Active Choice (17.2%), Reserved for You (16.6%) and Stand Proud (14.5%) forms, and fewer bookings/referrals were made among those who submitted a Foot-in-the-Door form (11.3%) (Figure 2C). The adjusted regression analysis (Table 2, Model 4) found that users submitting on the Foot-in-the-Door form were *less* likely to be booked/referred compared to SOC (-5 percentage points, p<0.05). None of the other relationships were different at p<0.05.

### Circumcised

A total of 48 individuals (1.2%) in our sub-sample were recorded in clinic records as having been circumcised. Compared to the SOC form, a greater proportion submitting on all other forms were circumcised (Figure 2D), but differences were not statistically significant in the adjusted regression analysis (Table 2, Model 6). Regression results for the secondary analysis were substantively similar in logistic regression models (Supplementary Table S6) and with the inclusion of a control variable to account for someone submitting multiple forms (Supplementary Table S7).

## Discussion

This study assessed whether messages framed using behavioural economics principles could encourage more MoyaApp users to submit their details for VMMC. Users viewing the form that leveraged the foot-in-the-door technique were significantly more likely to submit a form compared to those viewing the SOC form. Those who viewed the Active Choice and Reserved for You forms were significantly less likely to submit a form for further engagement. Studies utilising the foot-in-the-door technique in HIV prevention in low- and middle-income countries are limited. One study in the USA showed that creating opportunities for participants to engage in HIV prevention activities ranging from the lighter or less time-consuming activities to progressively heavier or more time-consuming activities (the foot-in-the-door concept)^19^ was effective: perusing an HIV prevention brochure increased watching a 10-minute video on HIV prevention, and watching the video increased likelihood of engaging in a counselling session^19^.

Research in other contexts and experimental conditions have shown mixed results and indicate that the foot-in-the-door effect may be influenced by various psychological processes (e.g., self-perception and intention), and that the strength of these processes can either enhance or reduce the effect^29,30^. Influencing attitudes, social norms and perceived behavioural control can strengthen the intention to carry out health behaviours^16^, in this case acting on the intention to undergo medical circumcision.

While the Foot-in-the-Door form did better at encouraging form submissions, it did worse in terms of successful bookings/referrals for an appointment. In contrast, messaging based on Reserved for You and Active Choice principles performed marginally better than the SOC form on bookings for VMMC (although differences were not statistically significant) despite having generated relatively fewer initial form submissions. Circumcision outcomes for MoyaApp users submitting VMMC forms were not different between the intervention and SOC forms.

The reasons for the differences across outcomes are unknown. It is possible that the foot-in-the-door technique encouraged more submissions because it lowered the sense of initial commitment. However, people who submitted the Foot-in-the-Door form may have also been less interested in VMMC overall than those submitting other forms. It is also possible that an increase in submissions from a successful form resulted in increased pressure on the call centre due to greater volumes of people to contact. It is not known whether the additional volume of pending calls could have influenced the time spent, quality and the extent to which questions were addressed in discussions the call centre agents had with potential clients. Since more bookings were made on the Stand Proud, Reserved for You and Active Choice forms, combining aspects of these forms with Foot-in-the-Door may have the potential for an enhanced effect and should be investigated further.

Following form submission, call centre engagement with people was generalised to the standard programmatic approach to bookings/referrals and did not consider the specific messaging per MoyaApp VMMC form. It would be important to explore whether an intervention at the stage of call centre contact that continues the behaviourally informed dialogue started in the form, could be effective in motivating men to overcome the intention-action gap to being circumcised. Behaviourally informed scripts could be used by call centre agents to align discussions with the behavioural economics principles applied in the MoyaApp VMMC form. Such scripts should take into consideration the stage at which a man may be in contemplating medical circumcision and specific questions that have frequently been raised before. A VMMC framework using market research and a person-centred approach in Zambia and Zimbabwe identified three stages (relate, anticipate, relieve) in understanding steps towards medical circumcision^13^. Each stage includes psychological processes that men may grapple with in following through on the intention to go for medical circumcision^13^. The framework, adaptable to local contexts, could be used to enhance the intervention forms and inform scripts that call centre counsellors use to encourage acting on the intention to circumcise, while controlling for system-related influences^13^.

The results of this study should be interpreted alongside its limitations. Since only one VMMC form was viewable in the MoyaApp at a time, it was not possible to randomise users to study arms. Furthermore, there was a deviation in the predetermined sequence of the forms appearing in the MoyaApp, resulting in the same form appearing on two consecutive Fridays at two points in the study. While this might have impacted form submissions, any bias was likely mitigated by controlling for temporal influences on form submissions in the regression analyses. A proportion of MoyaApp users who submitted forms were not attempted to be contacted by the call centre, which could have introduced bias in successful contact, booking/referral and circumcision outcomes. Submissions on the SOC form were more likely to be excluded from follow-up contact by the call centre than submissions on each of the intervention forms. It is unclear whether this might have biased results. One possibility is that the SOC group received better attention from the call centre (i.e., relatively fewer people to call may have given the call centre more time to make calls and speak with people) which could have biased results towards the SOC.

The study was limited to data from MoyaApp users and VMMC programme data for people 18 years and older. A recent study indicated that younger men (below 20 years of age) often access medical circumcision more than older men (20-34 years)^31^. Younger men are an important target population for medical circumcision and it is important to assess the impact of message framing among these individuals.

## Conclusions

Message framing using behavioural economics principles can be used in the HIV context to nudge men to engage with health services. The Foot-in-the-Door intervention was effective in increasing interest in medical circumcision. However, more work is needed on motivating the continuation of this interest and conversion to acting on the intention to VMMC downstream of the initial interest. Incorporating behavioural economics strategies in targeted interventions that are low cost, scalable and take a person-centred approach can be effective in motivating engagement in care. The use of technology provides a low-cost and convenient platform to implement interventions to encourage uptake of health services. Future research on digital message framing interventions to increase VMMC should include younger men, who may engage with technology differently and are more likely than older men to get circumcised. Learnings from this work could also be applied and evaluated in other areas of HIV prevention and treatment.

## Data Availability

All data produced in the present study are available upon reasonable request to the authors

## Acknowledgements

We would like to thank all the data service providers for granting access to the data including our study staff and Right to Care staff for their support and contribution to the success of the study. A special gratitude goes to the core Right to Care VMMC Programme staff (Khumbulani Moyo, Sizwe Hlongwane, Nelson Igaba, Motshana Phohole).

## Disclosures and Disclaimers Funding

This analysis was funded by the Bill and Melinda Gates Foundation (INV-008318). The contents are the responsibility of the authors and do not necessarily reflect the views of the sponsor. LL was partially supported by the National Institute of Mental Health of the National Institutes of Health under grant number K01MH119923. The funders had no role in the study design, collection, analysis and interpretation of the data, in manuscript preparation or the decision to publish.

We wish to acknowledge the support from the University of California, San Francisco’s International Traineeships in AIDS Prevention Studies (ITAPS), U.S. NIMH, R25MH123256 and IAVI. This work was assisted in part by a CFAR/ARI HIV Research Boost Award from the UCSF AIDS Research Institute. The content is solely the responsibility of the authors and does not necessarily represent the official views of the NIH, NIMH, FIC or other funding institutes.

## Supplemental Digital Content

Supplementary tables S1-S7 included.

