## Supplemental files for "Evaluation of four interventions using behavioural economics insights to increase demand for voluntary medical male circumcision in South Africa through the MoyaApp: A quasi-experimental study"

**Supplementary Materials**

**Table S1. Sequencing of VMMC study forms on the MoyaApp**

| | **Monday** | **Wednesday** | **Friday** |
|---|---|---|---|
| **W1** | Stand Proud (SP) | Active Choice (AC) | Reserved for You (RFY) |
| **W2** | Foot in the Door (FITD) | Standard of Care (SOC) | Stand Proud (SP) |
| **W3** | Active Choice (AC) | Reserved for You (RFY) | Foot in the Door (FITD) |
| **W4** | Standard of Care (SOC) | Stand Proud (SP) | Active Choice (AC) |
| **W5** | Reserved for You (RFY) | Foot in the Door (FITD) | Standard of Care (SOC) |
| **W6** | Stand Proud (SP) | Active Choice (AC) | Reserved for You (RFY) |
| **W7** | Foot in the Door (FITD) | Standard of Care (SOC) | **Reserved for You (RFY)*** |
| **W8** | Active Choice (AC) | Reserved for You (RFY) | Foot in the Door (FITD) |
| **W9** | Standard of Care (SOC) | Stand Proud (SP) | Active Choice (AC) |
| **W10** | Reserved for You (RFY) | Foot in the Door (FITD) | Standard of Care (SOC) |
| **W11** | Stand Proud (SP) | Active Choice (AC) | **Stand Proud (SP)*** |
| **W12** | Foot in the Door (FITD) | Standard of Care (SOC) | Stand Proud (SP) |
| **W13** | Active Choice (AC) | Reserved for You (RFY) | Foot in the Door (FITD) |
| **W14** | Standard of Care (SOC) | Stand Proud (SP) | Active Choice (AC) |
| **W15** | Reserved for You (RFY) | Foot in the Door (FITD) | Standard of Care (SOC) |

*Deviation in form rotation in W7 compensated for in W11

**Table S2 Ordinary least squares regression models assessing the association between VMMC forms submitted across five study groups by adults who viewed the forms during August and November 2022 in South Africa (N=118,337)**

| | | | Submitted Forms | |
|---|---|---|---|---|
| | | | Unadjusted (Model 1) | Adjusted (Model 2) |
| | N | % Submissions | **β** [95% CI] | **β** [95% CI] |
| Study groups (ref: Standard of Care, SOC) | 24,459 | 6.3 | | |
| Foot-in-the-Door | 22,351 | 7.4 | 0.012*** [0.004 - 0.019] | 0.013*** [0.009 - 0.017] |
| Stand Proud | 23,195 | 5.8 | -0.005 [-0.012 - 0.002] | -0.002 [-0.008 - 0.003] |
| Reserved for You | 24,178 | 5.6 | -0.007* [-0.014 - 0.000] | -0.005** [-0.009 - -0.001] |
| Active Choice | 24,154 | 5.0 | -0.013*** [-0.019 - -0.006] | -0.011*** [-0.017 - -0.006] |
| Day of the week controls (ref Monday) | | | | |
| Wednesday | | | 0.004*** | [0.001 - 0.007] |
| Friday | | | 0.002 | [-0.001 - 0.005] |
| Study week controls (ref week 1) | | | | |
| Week 2 | | | | 0.001] [-0.007 - 0.009 |
| Week 3 | | | | 0.013*** [0.006 - 0.019] |
| Week 4 | | | | 0.012*** [0.006 - 0.019] |
| Week 5 | | | | 0.012*** [0.007 - 0.018] |
| Week 6 | | | | 0.015*** [0.011 - 0.018] |
| Week 7 | | | | 0.011*** [0.007 - 0.015] |
| Week 8 | | | | 0.012*** [0.009 - 0.015] |
| Week 9 | | | | 0.016*** [0.010 - 0.023] |
| Week 10 | | | | 0.021*** [0.017 - 0.024] |
| Week 11 | | | | 0.013*** [0.009 - 0.017] |
| Week 12 | | | | 0.019*** [0.014 - 0.024] |
| Week 13 | | | | 0.016*** [0.009 - 0.023] |
| Week 14 | | | | 0.016*** [0.006 - 0.025] |

| | | |
|---|---|---|
| Week 15 | | 0.016*** |
| | | [0.008 - 0.024] |
| Observations | 118,337 | 118,337 |
| R-squared | 0.001 | 0.002 |

* p<0.1, ** p<0.05, *** p<0.01

**Table S3 Logistic regression models assessing the association between VMMC forms submitted across five study groups by adults who viewed the forms during August and November 2022 in South Africa (N=118,337)**

| | | | Submitted Forms | |
|---|---|---|---|---|
| | | | Unadjusted (Model 1) | Adjusted (Model 2) |
| | N | % Submissions | Odds ratio [95% CI] | Odds ratio [95% CI] |
| Study groups (ref: Standard of Care, SOC) | 24,459 | 6.3 | | |
| Foot-in-the-Door | 22,351 | 7.4 | 1.204*** [1.071 - 1.353] | 1.220*** [1.145 - 1.300] |
| Stand Proud | 23,195 | 5.8 | 0.915 [0.810 - 1.033] | 0.956 [0.871 - 1.049] |
| Reserved for You | 24,178 | 5.6 | 0.884* [0.781 - 1.000] | 0.913** [0.852 - 0.980] |
| Active Choice | 24,154 | 5 .0 | 0.789*** [0.701 - 0.887] | 0.806*** [0.729 - 0.892] |
| Day of the week controls (ref Monday) | | | | |
| Wednesday | | | | 1.070** [1.016 - 1.128] |
| Friday | | | | 1.031 [0.979 - 1.086] |
| Study week controls (ref week 1) | | | | |
| Week 2 | | | | 1.057 [0.911 - 1.226] |
| Week 3 | | | | 1.317*** [1.176 - 1.475] |
| Week 4 | | | | 1.312*** [1.161 - 1.482] |
| Week 5 | | | | 1.302*** [1.179 - 1.439] |
| Week 6 | | | | 1.369*** [1.265 - 1.482] |
| Week 7 | | | | 1.278*** [1.189 - 1.375] |
| Week 8 | | | | 1.306*** [1.226 - 1.392] |
| Week 9 | | | | 1.410*** [1.250 - 1.589] |
| Week 10 | | | | 1.491*** [1.384 - 1.606] |
| Week 11 | | | | 1.330*** [1.221 - 1.450] |
| Week 12 | | | | 1.460*** [1.338 - 1.594] |
| Week 13 | | | | 1.399*** |

| | | |
|---|---|---|
| | | [1.237 - 1.581] |
| Week 14 | | 1.390*** |
| | | [1.166 - 1.657] |
| Week 15 | | 1.391*** |
| | | [1.221 - 1.585] |
| Observations | 118,337 | 118,337 |

* p<0.1, ** p<0.05, *** p<0.01

**Table S4 Ordinary least squares regression on factors associated with not being included on the contact list**

| | | Contacts Attempted | |
|---|---|---|---|
| | N | % Contacts Attempted | Adjusted (Model 2) β [95% CI] |
| Study groups (ref: Standard of Care, SOC) | 1,433 | 51.8 | |
| Foot-in-the-Door | 1,576 | 62.1 | 0.129*** [0.093 - 0.166] |
| Stand Proud | 1,262 | 70.9 | 0.181*** [0.140 - 0.223] |
| Reserved for You | 1,261 | 64.4 | 0.109*** [0.069 - 0.148] |
| Active Choice | 1,120 | 64.8 | 0.117*** [0.075 - 0.159] |
| Day of the week controls (ref: Monday) | | | |
| Wednesday | | | -0.029** [-0.058 - -0.000] |
| Friday | | | -0.039*** [-0.066 - -0.011] |
| Study week controls (ref 1) | | | |
| Week 2 | | | -0.113*** [-0.187 - -0.039] |
| Week 3 | | | 0.052 [-0.019 - 0.123] |
| Week 4 | | | -0.072** [-0.142 - -0.002] |
| Week 5 | | | 0.013 [-0.058 - 0.085] |
| Week 6 | | | 0.076** [0.007 - 0.145] |
| Week 7 | | | 0.009 [-0.060 - 0.078] |
| Week 8 | | | -0.244*** [-0.314 - -0.174] |
| Week 9 | | | 0.111*** [0.037 - 0.186] |
| Week 10 | | | -0.066* [-0.135 - 0.004] |
| Week 11 | | | 0.050 [-0.021 - 0.121] |
| Week 12 | | | -0.195*** [-0.266 - -0.124] |
| Week 13 | | | -0.103*** [-0.177 - -0.029] |
| Week 14 | | | -0.045 [-0.119 - 0.030] |
| Week 15 | | | 0.091** [0.018 - 0.165] |
| Observations | | | 6,652 |
| R-squared | | | 0.060 |

* p<0.1, ** p<0.05, *** p<0.01

A total of 2,498 out of 6652 submitted forms were not included in the contact list due to an administrative error  (i.e., no contact attempts were made based on these form)

**Table S5 Ordinary least squares regression models of the impact of the study forms on secondary study outcomes: successful contact by the call centre, booking/referral complete, circumcision complete**

| | N | % Contacted | **Contacted** Unadjusted (Model 1) **β** [95% CI] | Adjusted (Model 2) **β** [95% CI] | % Booked/Referred | **Booked/Referred** Unadjusted (Model 3) **β** [95% CI] | Adjusted (Model 4) **β** [95% CI] | % Circumcised | **Circumcised** Unadjusted (Model 5) **β** [95% CI] | Adjusted (Model 6) **β** [95% CI] |
|---|---|---|---|---|---|---|---|---|---|---|
| Study groups (ref: Standard of Care, SOC) | 743 | 54.9% | | | 14.4% | | | 0.7% | | |
| Foot-in-the-Door | 978 | 56.3% | 0.014 [-0.053 - 0.081] | -0.016 [-0.068 - 0.037] | 11.3% | -0.031 [-0.073 - 0.012] | -0.050** [-0.092 - -0.009] | 1.1% | 0.005 [-0.004 - 0.013] | 0.002 [-0.007 - 0.011] |
| Stand Proud | 895 | 47.3% | -0.076** [-0.145 - -0.008] | -0.038 [-0.091 - 0.016] | 14.5% | 0.001 [-0.041 - 0.043] | -0.031 [-0.081 - 0.018] | 1.1% | 0.004 [-0.005 - 0.014] | 0 [-0.012 - 0.011] |
| Reserved for You | 812 | 58.1% | 0.032 [-0.048 - 0.112] | 0.040* [-0.007 - 0.087] | 16.6% | 0.022 [-0.029 - 0.073] | 0.01 [-0.032 - 0.053] | 0.9% | 0.002 [-0.007 - 0.011] | 0.001 [-0.008 - 0.011] |
| Active Choice | 726 | 55.0% | 0 [-0.066 - 0.067] | 0.036 [-0.021 - 0.093] | 17.2% | 0.028 [-0.016 - 0.073] | 0.011 [-0.038 - 0.059] | 2.1% | 0.014** [0.000 - 0.027] | 0.012* [-0.000 - 0.025] |
| Day of the week controls (ref: Monday) | | | | | | | | | | |
| Wednesday | | | | 0.021 [-0.011 - 0.053] | | | 0.000 [-0.032 - 0.033] | | | -0.005 [-0.012 - 0.003] |
| Friday | | | | -0.037** [-0.073 - -0.000] | | | -0.019 [-0.051 - 0.013] | | | -0.003 [-0.011 - 0.006] |

| | | | |
|---|---|---|---|
| Study week controls (ref 1) | | | |
| Week 2 | 0.045<br>[-0.095 - 0.185] | 0.002<br>[-0.135 - 0.138] | 0.001<br>[-0.030 - 0.032] |
| Week 3 | 0.107***<br>[0.045 - 0.168] | -0.026<br>[-0.120 - 0.068] | -0.004<br>[-0.028 - 0.020] |
| Week 4 | 0.053<br>[-0.025 - 0.130] | -0.002<br>[-0.103 - 0.098] | -0.004<br>[-0.029 - 0.021] |
| Week 5 | 0.124***<br>[0.057 - 0.192] | -0.046<br>[-0.157 - 0.066] | -0.002<br>[-0.028 - 0.024] |
| Week 6 | 0.077***<br>[0.021 - 0.134] | -0.023<br>[-0.113 - 0.066] | -0.013<br>[-0.037 - 0.011] |
| Week 7 | 0.140***<br>[0.048 - 0.231] | -0.041<br>[-0.142 - 0.060] | -0.005<br>[-0.030 - 0.020] |
| Week 8 | -0.072<br>[-0.161 - 0.017] | -0.057<br>[-0.155 - 0.042] | -0.003<br>[-0.029 - 0.023] |
| Week 9 | 0.052<br>[-0.013 - 0.117] | -0.062<br>[-0.164 - 0.039] | -0.005<br>[-0.030 - 0.020] |
| Week 10 | 0.143***<br>[0.069 - 0.218] | -0.030<br>[-0.130 - 0.070] | -0.003<br>[-0.028 - 0.021] |
| Week 11 | 0.051*<br>[-0.010 - 0.113] | -0.010<br>[-0.111 - 0.091] | 0.010<br>[-0.014 - 0.033] |
| Week 12 | 0.109*** | 0.009 | -0.005 |

| | | | | | | |
|---|---|---|---|---|---|---|
| | | [0.043 - 0.175] | | [-0.080 - 0.097] | | [-0.031 - 0.021] |
| Week 13 | | 0.252*** | | 0.018 | | 0.009 |
| | | [0.187 - 0.316] | | [-0.100 - 0.136] | | [-0.025 - 0.043] |
| Week 14 | | 0.112* | | -0.045 | | -0.008 |
| | | [-0.001 - 0.225] | | [-0.142 - 0.052] | | [-0.036 - 0.020] |
| Week 15 | | 0.090** | | -0.091* | | -0.009 |
| | | [0.004 - 0.176] | | [-0.183 - 0.002] | | [-0.033 - 0.015] |
| Observations | 4,154 | 4,154 | 4,154 | 4,154 | 4,154 | 4,154 |
| R-squared | 0.006 | 0.021 | 0.004 | 0.009 | 0.002 | 0.005 |

* p<0.1, ** p<0.05, *** p<0.01

**Table S6 Logistic regression models of VMMC engagement in the MoyaApp (dependent variables: successful contact, booking/referral, circumcision) (N=4,154)**

| | | Contacted | | Booked/Referred | | Circumcised | |
|---|---|---|---|---|---|---|---|
| | N | % Contacted | Adjusted (Model 1) Odds ratio [95% CI] | % Booked/Referred | Adjusted (Model 2) Odds ratio [95% CI] | % Circumcised | Adjusted (Model 3) Odds ratio [95% CI] |
| Study groups (ref: Standard of Care, SOC) | 743 | 54.9% | | 14.4% | | 0.7% | |
| Foot-in-the-Door | 978 | 56.3% | 0.924 [0.750 - 1.139] | 11.3% | 0.642*** [0.462 - 0.892] | 1.1% | 1.333 [0.473 - 3.755] |
| Stand Proud | 895 | 47.3% | 0.825* [0.669 - 1.018] | 14.5% | 0.737* [0.520 - 1.044] | 1.1% | 0.910 [0.253 - 3.267] |
| Reserved for You | 812 | 58.1% | 1.169* [0.971 - 1.407] | 16.6% | 1.062 [0.769 - 1.467] | 0.9% | 1.145 [0.361 - 3.637] |
| Active Choice | 726 | 55.0% | 1.100 [0.883 - 1.370] | 17.2% | 0.994 [0.699 - 1.414] | 2.1% | 2.263 [0.815 - 6.284] |
| Day of the week controls (ref: Monday) | | | | | | | |
| Wednesday | | | 1.096 [0.960 - 1.251] | | 0.979 [0.776 - 1.237] | | 0.638 [0.326 - 1.248] |
| Friday | | | 0.861** | | 0.866 | | 0.847 |

| | | | |
|---|---|---|---|
| | [0.747 - 0.993] | [0.675 - 1.112] | [0.456 - 1.575] |
| Study week controls (ref Week 1) | | | |
| Week 2 | 1.209<br>[0.695 - 2.105] | 1.031<br>[0.386 - 2.754] | 1.059<br>[0.136 - 8.226] |
| Week 3 | 1.573***<br>[1.209 - 2.046] | 0.823<br>[0.425 - 1.595] | 0.664<br>[0.133 - 3.303] |
| Week 4 | 1.266<br>[0.919 - 1.746] | 1.025<br>[0.512 - 2.049] | 0.754<br>[0.119 - 4.788] |
| Week 5 | 1.651***<br>[1.240 - 2.199] | 0.687<br>[0.296 - 1.596] | 0.791<br>[0.117 - 5.337] |
| Week 6 | 1.394***<br>[1.098 - 1.769] | 0.868<br>[0.468 - 1.611] | 0.183<br>[0.019 - 1.800] |
| Week 7 | 1.803***<br>[1.238 - 2.626] | 0.756<br>[0.369 - 1.547] | 0.627<br>[0.095 - 4.149] |
| Week 8 | 0.761<br>[0.526 - 1.101] | 0.672<br>[0.334 - 1.350] | 0.758<br>[0.141 - 4.085] |
| Week 9 | 1.233<br>[0.951 - 1.597] | 0.605<br>[0.297 - 1.234] | 0.624<br>[0.105 - 3.713] |
| Week 10 | 1.813*** | 0.807 | 0.750 |

| | | | |
|---|---|---|---|
| | [1.328 - 2.475] | [0.413 - 1.580] | [0.129 - 4.358] |
| Week 11 | 1.310** | 1.028 | 2.052 |
| | [1.009 - 1.701] | [0.509 - 2.075] | [0.470 - 8.948] |
| Week 12 | 1.645*** | 1.190 | 0.721 |
| | [1.247 - 2.168] | [0.645 - 2.193] | [0.086 - 6.013] |
| Week 13 | 3.097*** | 1.235 | 1.778 |
| | [2.295 - 4.179] | [0.570 - 2.678] | [0.288 - 10.957] |
| Week 14 | 1.672** | 0.787 | 0.618 |
| | [1.084 - 2.578] | [0.406 - 1.525] | [0.073 - 5.218] |
| Week 15 | 1.520** | 0.474** | 0.326 |
| | [1.072 - 2.155] | [0.254 - 0.886] | [0.030 - 3.537] |
| Multiple submissions (ref single submission) | 2.610*** | 3.206*** | 4.798*** |
| | [1.968 - 3.462] | [2.402 - 4.279] | [2.619 - 8.789] |
| Observations | 4,154 | 4,154 | 4,154 |

* p<0.1, ** p<0.05, *** p<0.01

**Table S7 Ordinary least squares regression models of VMMC engagement in the MoyaApp (dependent variables: successful contact, booking/referral, circumcision) (N=4,154) with controls for multiple submitters**

| | | **Contacted** | | **Booked/Referred** | | **Circumcised** | |
|---|---|---|---|---|---|---|---|
| | N | % Contacted | Adjusted (Model 1) **β** [95% CI] | % Booked/Referred | Adjusted (Model 2) **β** [95% CI] | % Circumcised | Adjusted (Model 3) **β** [95% CI] |
| Study groups (ref: Standard of Care, SOC) | 743 | 54.9% | | 14.4% | | 0.7% | |
| Foot-in-the-Door | 978 | 56.3% | -0.017 [-0.069 - 0.035] | 11.3% | -0.052*** [-0.090 - -0.014] | 1.1% | 0.002 [-0.007 - 0.011] |
| Stand Proud | 895 | 47.3% | -0.047* [-0.100 - 0.006] | 14.5% | -0.040* [-0.086 - 0.007] | 1.1% | -0.002 [-0.013 - 0.010] |
| Reserved for You | 812 | 58.1% | 0.036 [-0.010 - 0.081] | 16.6% | 0.007 [-0.035 - 0.049] | 0.9% | 0.001 [-0.009 - 0.011] |
| Active Choice | 726 | 55.0% | 0.023 [-0.032 - 0.078] | 17.2% | -0.001 [-0.048 - 0.046] | 2.1% | 0.011* [-0.002 - 0.023] |
| Day of the week controls (ref: Monday) | | | | | | | |
| Wednesday | | | 0.022 [-0.011 - 0.054] | | 0.001 [-0.030 - 0.032] | | -0.005 [-0.012 - 0.003] |
| Friday | | | -0.035* [-0.071 - 0.000] | | -0.018 [-0.049 - 0.014] | | -0.002 [-0.011 - 0.006] |
| Study week controls (ref 1) | | | | | | | |
| Week 2 | | | 0.044 [-0.093 - 0.182] | | 0.001 [-0.132 - 0.134] | | 0.001 [-0.030 - 0.032] |
| Week 3 | | | 0.108*** [0.044 - 0.172] | | -0.025 [-0.122 - 0.072] | | -0.004 [-0.028 - 0.020] |
| Week 4 | | | 0.057 | | 0.002 | | -0.003 |

| | | | |
|---|---|---|---|
| | [-0.023 - 0.137] | [-0.102 - 0.105] | [-0.029 - 0.023] |
| Week 5 | 0.121*** | -0.049 | -0.003 |
| | [0.050 - 0.192] | [-0.164 - 0.066] | [-0.029 - 0.024] |
| Week 6 | 0.081*** | -0.020 | -0.013 |
| | [0.022 - 0.140] | [-0.113 - 0.072] | [-0.037 - 0.012] |
| Week 7 | 0.143*** | -0.039 | -0.004 |
| | [0.049 - 0.236] | [-0.141 - 0.064] | [-0.030 - 0.021] |
| Week 8 | -0.066 | -0.052 | -0.002 |
| | [-0.157 - 0.024] | [-0.153 - 0.049] | [-0.028 - 0.024] |
| Week 9 | 0.050 | -0.064 | -0.005 |
| | [-0.014 - 0.114] | [-0.165 - 0.037] | [-0.030 - 0.020] |
| Week 10 | 0.143*** | -0.030 | -0.003 |
| | [0.067 - 0.219] | [-0.130 - 0.071] | [-0.028 - 0.022] |
| Week 11 | 0.064** | 0.002 | 0.012 |
| | [0.000 - 0.129] | [-0.100 - 0.104] | [-0.012 - 0.036] |
| Week 12 | 0.119*** | 0.018 | -0.003 |
| | [0.051 - 0.188] | [-0.073 - 0.109] | [-0.029 - 0.023] |
| Week 13 | 0.262*** | 0.027 | 0.011 |
| | [0.195 - 0.329] | [-0.094 - 0.148] | [-0.024 - 0.046] |
| Week 14 | 0.125** | -0.033 | -0.006 |
| | [0.016 - 0.234] | [-0.131 - 0.064] | [-0.033 - 0.022] |
| Week 15 | 0.101** | -0.081* | -0.007 |
| | [0.014 - 0.188] | [-0.176 - 0.014] | [-0.032 - 0.017] |
| Multiple submissions (ref single submission) | 0.215*** | 0.194*** | 0.032*** |
| | [0.157 - 0.273] | [0.134 - 0.254] | [0.010 - 0.055] |
| Observations | 4,154 | 4,154 | 4,154 |
| R-squared | 0.034 | 0.030 | 0.011 |

* p<0.1, ** p<0.05, *** p<0.01
